# Large language models for conducting systematic reviews: on the rise, but not yet ready for use – a scoping review

**DOI:** 10.1101/2024.12.19.24319326

**Authors:** Judith-Lisa Lieberum, Markus Töws, Maria-Inti Metzendorf, Felix Heilmeyer, Waldemar Siemens, Christian Haverkamp, Daniel Böhringer, Joerg J. Meerpohl, Angelika Eisele-Metzger

## Abstract

**Background:** Machine learning (ML) promises versatile help in the creation of systematic reviews (SRs). Recently, further developments in the form of large language models (LLMs) and their application in SR conduct attracted attention.

**Objective:** To provide an overview of ML and specifically LLM applications in SR conduct in health research.

**Study design:** We systematically searched MEDLINE, Web of Science, IEEEXplore, ACM Digital Library, Europe PMC (preprints), Google Scholar, and conducted an additional hand search (last search: 26 February 2024). We included scientific articles in English or German, published from April 2021 onwards, building upon the results of a mapping review with a related research question. Two reviewers independently screened studies for eligibility; after piloting, one reviewer extracted data, checked by another.

**Results:** Our database search yielded 8054 hits, and we identified 33 articles from our hand search. Of the 196 included reports, 159 described more traditional ML techniques, 37 focused on LLMs. LLM approaches covered 10 of 13 defined SR steps, most frequently literature search (n=15, 41%), study selection (n=14, 38%), and data extraction (n=11, 30%). The mostly recurring LLM was GPT (n=33, 89%). Validation studies were predominant (n=21, 57%). In half of the studies, authors evaluated LLM use as promising (n=20, 54%), one quarter as neutral (n=9, 24%) and one fifth as non-promising (n=8, 22%).

**Conclusions:** Although LLMs show promise in supporting SR creation, fully established or validated applications are often lacking. The rapid increase in research on LLMs for evidence synthesis production highlights their growing relevance.

**HIGHLIGHTS:**

- Machine learning (ML) offers promising support for systematic review (SR) creation.
- GPT was the most commonly used large language model (LLM) to support SR production.
- LLM application included 10 of 13 defined SR steps, most often literature search.
- Validation studies predominated, but fully established LLM applications are rare.
- LLM research for SR conduct is surging, highlighting the increasing relevance.

## INTRODUCTION

Systematic reviews (SRs) form the basis for evidence-based medicine (EBM) [1, 2] but are time and resource intensive, requiring several researchers [3]. Using artificial intelligence (AI) to assist with the different SR tasks offers a promising approach to save time and decrease personnel demands [4, 5].

Recently, a wide array of AI tools has evolved: Traditional machine learning (ML) relies on supervised or unsupervised algorithms for task-specific decisions [6–9]. Transformers like BERT (Bidirectional Encoder Representations from Transformers) [10] vastly improved semantical and contextual language processing. Generative large language models (LLMs) such as GPT (Generative Pretrained Transformer) [11], LLaMA (Large Language Model Meta AI) [12] or Claude [13] follow instructions in natural language without task-specific training [14],. These are built on decoder-only transformers and trained on vast amounts of textual data. Yet, their opaque architecture raises risks like harmful responses or misinformation [14–16]. Currently, LLMs are extensively tested in medicine [17] and health research [18].

Several ML-based tools already assist SR conduction in health research, including ASReview for screening [19] and DistillerSR for various SR steps [20]. In a mapping review based on an April-May 2021 systematic search, Cierco Jimenez et al. [4] identified a broad range of ML tools that assist in SR performance. However, no LLM applications – perhaps holding even greater promise – could be identified at that time. Since then, LLM use in this context has risen significantly, aiding in tasks like formulating review questions [21], screening [22], or data extraction [23]. However, these approaches are still experimental and error prone. For instance, our recent attempts to assess risk of bias (RoB) with Cochrane’s risk of bias tool “RoB2” using Claude achieved limited success, far from replacing human reviewers [24].

This scoping review aims to provide an overview of recent approaches to facilitate SR conduct with the help of ML and LLMs in particular, highlighting the most promising strategies and forming a basis for future advancement and critical evaluation.

## METHODS

This scoping review was conducted based on the guidelines of the JBI (Joanna Briggs Institute) [25] and reported in line with the PRISMA-ScR statement (Preferred Reporting Items for Systematic reviews and Meta-Analyses extension for Scoping Reviews) [26].

### Protocol and registration

We registered the protocol for this scoping review on Open Science Framework (OSF) on March 4, 2024 (https://osf.io/asjm3). Changes from the protocol are listed in the supplement.

### Eligibility criteria

Eligibility criteria were defined following the PCC (population, concept, context) framework [25]; as common for methodological scoping reviews; limitations regarding population were not applicable. We included articles on ML applications (concept) in the context of conducting a SR in health research, published from 1 April 2021 onwards, building on the results of an earlier review on ML applications by Jimenez et al. who conducted their search in April and May 2021 [4]. Due to feasibility, inclusion was restricted to full scientific articles in English and German language, published in journals or on preprint servers. We considered support of any individual step or of the entire SR process. Specification of SR steps considered are listed in our protocol.

We excluded study or review protocols, preclinical literature, sources describing the application of ML tools specifically for guideline development and sources lacking detail regarding the tools and methods used or the area of application. Systematic, scoping, mapping or narrative reviews on ML approaches as well as surveys or guidance articles mentioning a range of approaches were included, but used for citation searching only, i.e. to check whether they cite further relevant articles that we may have missed with our systematic search.

### Information sources

We systematically searched MEDLINE via Ovid, Web of Science Core Collection (Science Citation Index Expanded, Social Science Citation Index, Emerging Sources Citation Index), IEEEXplore and ACM Digital Library (journals only), Europe PMC (preprints only) and Google Scholar (first 200 hits, sorted by relevance).

We conducted backward citation searching considering the review articles on ML approaches identified by our search as well as the review by Parisi and Sutton [27] on the role of ChatGPT in systematic literature searches published during our SR process. Additionally, we searched the Digital Evidence Synthesis Tool (DEST) Evaluations [28] and a range of literature collected from preliminary searches.

### Search

We established the search strategy based on text analysis of 38 known relevant records. Of those, 24 were indexed in PubMed, on which the search strategy for MEDLINE was developed by analyzing the most frequent MeSH terms with Yale MeSH Analyzer and conducting text word frequency analyses with PubReMiner. Additionally, we used parts of a search filter for Generative AI [29]. We subsequently revised the search strategy to capture the 14 records that were not indexed in PubMed and adapted the final search strategy to all sources. The date of search for all databases was 26 February 2024. The search results were deduplicated using Deduklick [30], resulting in 8054 records from database searches to be screened. Our final search strategies can be found in the supplement. In September 2024, we reviewed all preprint articles for a possible change of status to a peer-reviewed article, and used the journal article for data charting, if available.

### Selection of sources of evidence

The review team for source selection consisted of three reviewers (JLL, AEM and MT), all of who completed the initial pilot testing of 25 randomly selected sources: Using Covidence review software [30], two out of three reviewers independently reviewed eligibility of all records first on title and abstract basis, then as full text screening for the remaining articles. At each stage, we resolved discrepancies by discussion.

### Data charting process and data items

We made a distinction between our focus on LLMs and more traditional ML methods for data charting:

For LLM data, we developed a customized Covidence spreadsheet. We conducted a pilot testing with three randomly chosen sources extracted by two reviewers independently. After finalizing the spreadsheet and obtaining sufficient agreement during piloting, LLM data were extracted by one reviewer (JLL) and verified by a second (AEM). We extracted the types of LLM(s) used, the SR step(s) supported, location of the primary author, type of article, study design, methods in brief, key findings, limitations, funding, and conflicts of interest. We collected authors’ overall conclusions and rated them as “promising”, “neutral”, or “non-promising”, based on the statements of the study authors and considering the overall study results. For study design, we categorized studies as “validation study” if a defined reference standard was used and matching to this standard was calculated.

Data on more traditional ML approaches were extracted using an Excel 2016 spreadsheet: Review articles and articles on ML tools already included in Cierco Jimenez [4] were listed without further data charting. For new ML tools that had not been reported on in Cierco Jimenez, we charted the tools names (if available) and the SR step(s) supported. We classified the ML methods used into two categories: non-generative transformers (e.g. BERT, BART (Bidirectional and Auto-Regressive Transformers)) and classification methods using other techniques (e.g. K-Nearest Neighbors (KNN)). Classification was carried out by two reviewers experienced in the field (DB, FH). Ambiguities were collected and discussed.

Due to the nature of this review (scoping review), we did not critically appraise the underlying sources.

### Synthesis of results

We converted extracted LLM data from Covidence to Microsoft Excel 2016, performed statistical evaluations of frequencies with Excel and Stata (version 16.1), and generated graphical charts with R (version 4.4.2) and Microsoft PowerPoint 2016. Detailed LLM and ML data tables were uploaded to OSF (https://osf.io/vdsgb).

## RESULTS

Our systematic search of 6 databases yielded 11 323 hits from databases and further 33 records identified via others methods. Details of the selection process are depicted in the PRISMA flowchart (Figure 1). Finally, 196 studies were included, of which 83 focused on more traditional ML applications that had not previously been described in the mapping review by Cierco Jimenez et al., and 37 on LLM use.

**Figure 1:**
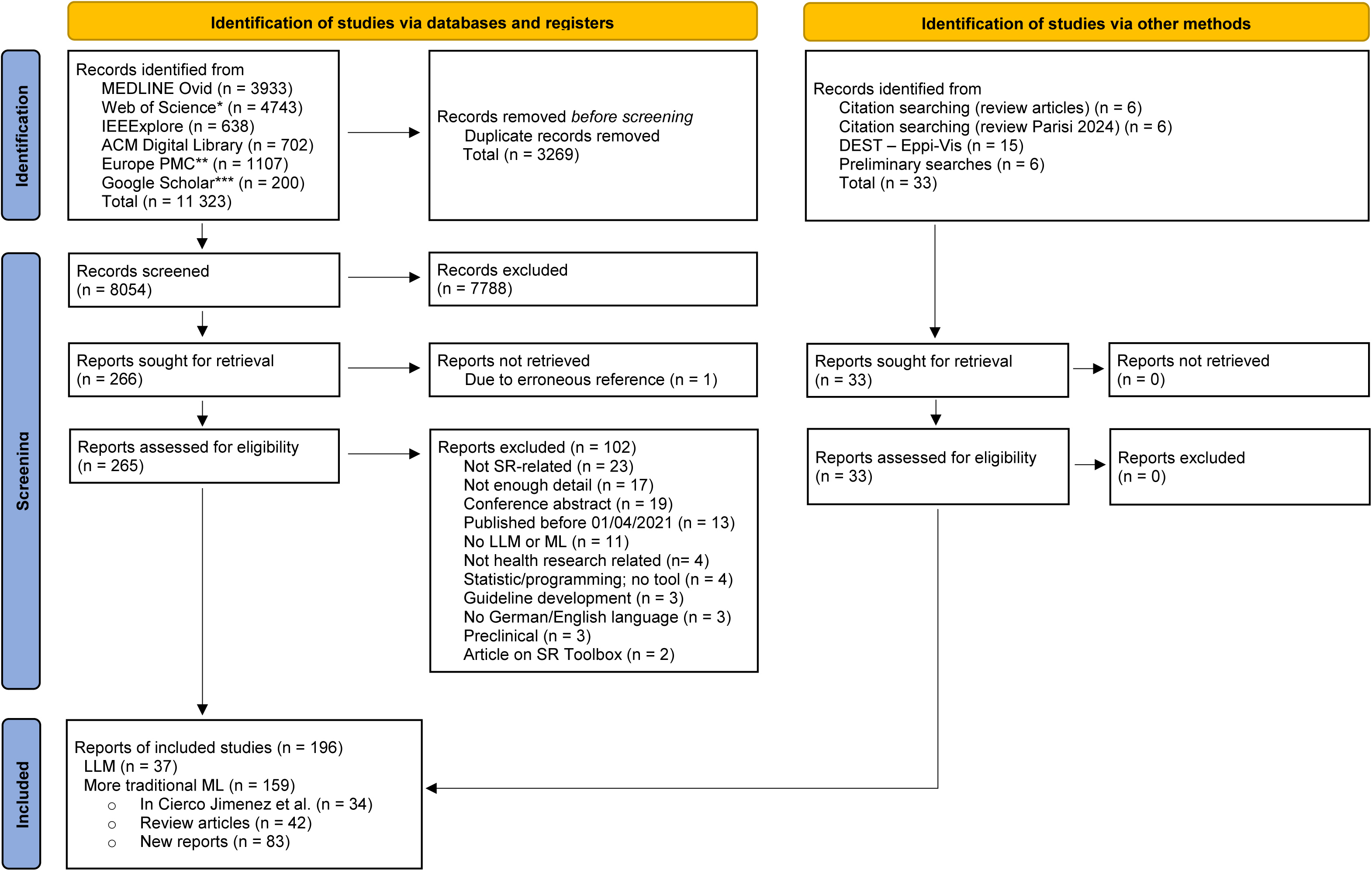
PRISMA flowchart describing the source of evidence retrieval and selection process of reports on the use of LLMs and ML in SR conduction in health research. * Web of Science Core Collection, ** Preprints only, *** First 200 hits sorted by relevance.

Of these reports on ML applications, we categorized 37% (n=30) as non-generative transformers, such as BERT or BART, and 59% (n=48) as other ML methods, e.g. KNN. Rarely, the technical basis of reports remained unclear (n=4, 5%). SR steps described by far most frequently were study selection (n=48, 47%), followed by search (n=21, 21%) and data extraction (n=12, 12%). Examples encompass COVIDScholar [31], a COVID-19 research aggregation and analysis platform supporting systematic search on COVID-19, based on natural language processing (NLP) techniques, or LiteRev [32], a NLP- and KNN-based automation tool facilitating search, data extraction, text retrieval and processing. An overview of reports on ML applications with differentiation of categories (new reports, review articles, and articles on tools included in Jimenez et al. in different sheets each) can be found on OSF (https://osf.io/vdsgb).

In the following, we concentrate on LLMs as the primary focus of our review (overview and characteristics in Table 1; data charting on OSF https://osf.io/vdsgb): The most frequent SR steps supported by LLMs were systematic literature search (n=15, in 41% of all 37 studies on LLM use), study selection including title/abstracts and full texts (n=14, 38%), and data extraction (n=11, 30%), followed by RoB assessment of primary studies (n=5, 14%), interpretation of findings (n=4, 11%), framing the SR question and inclusion criteria (n=3, 8%), and qualitative/narrative/descriptive summary of findings (n=3, 8%). LLMs were rarely used for writing PLS (n=1) and SR publications (n=2), or quantitative data analysis (n=2).

**Table 1:**
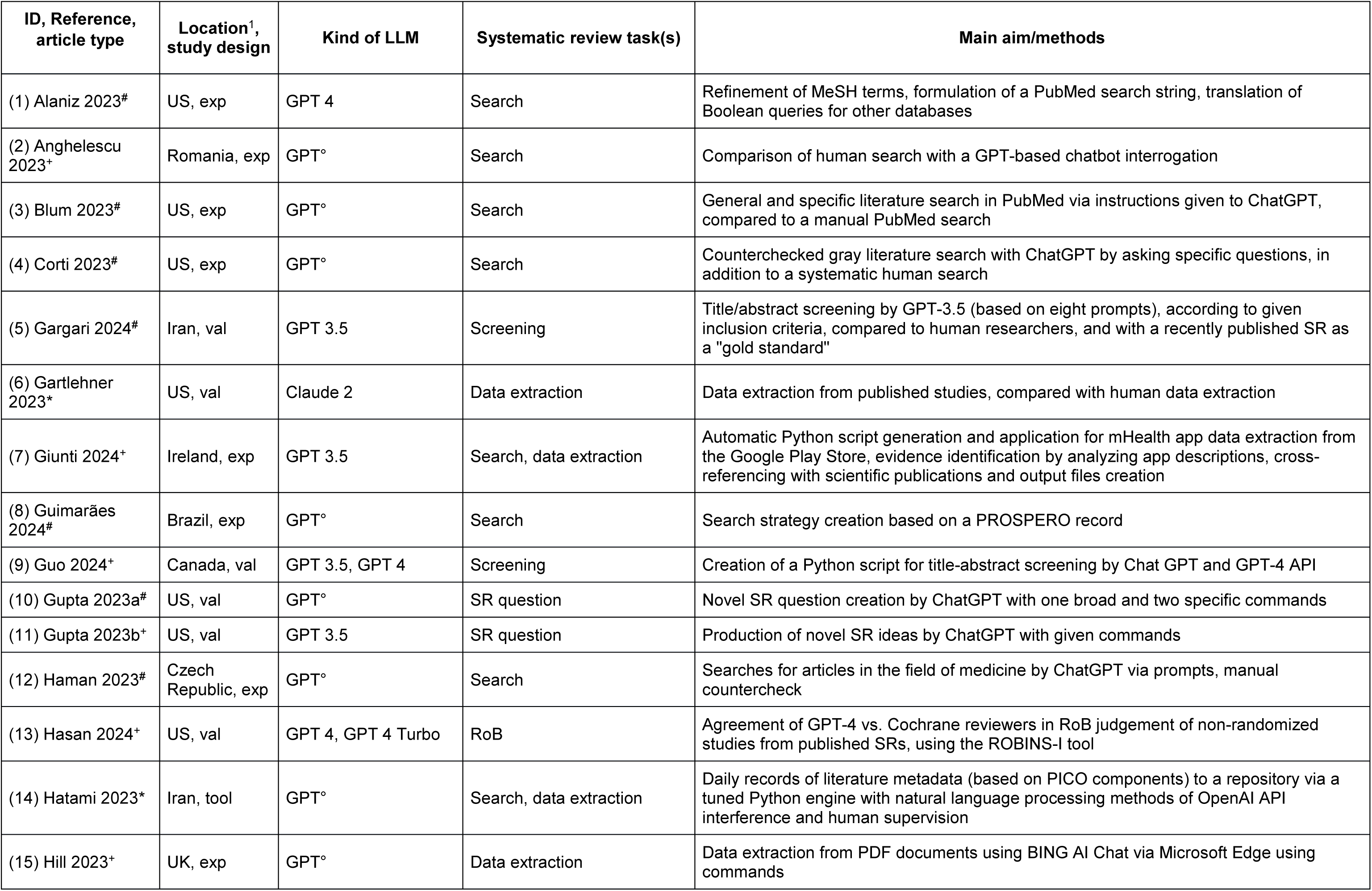
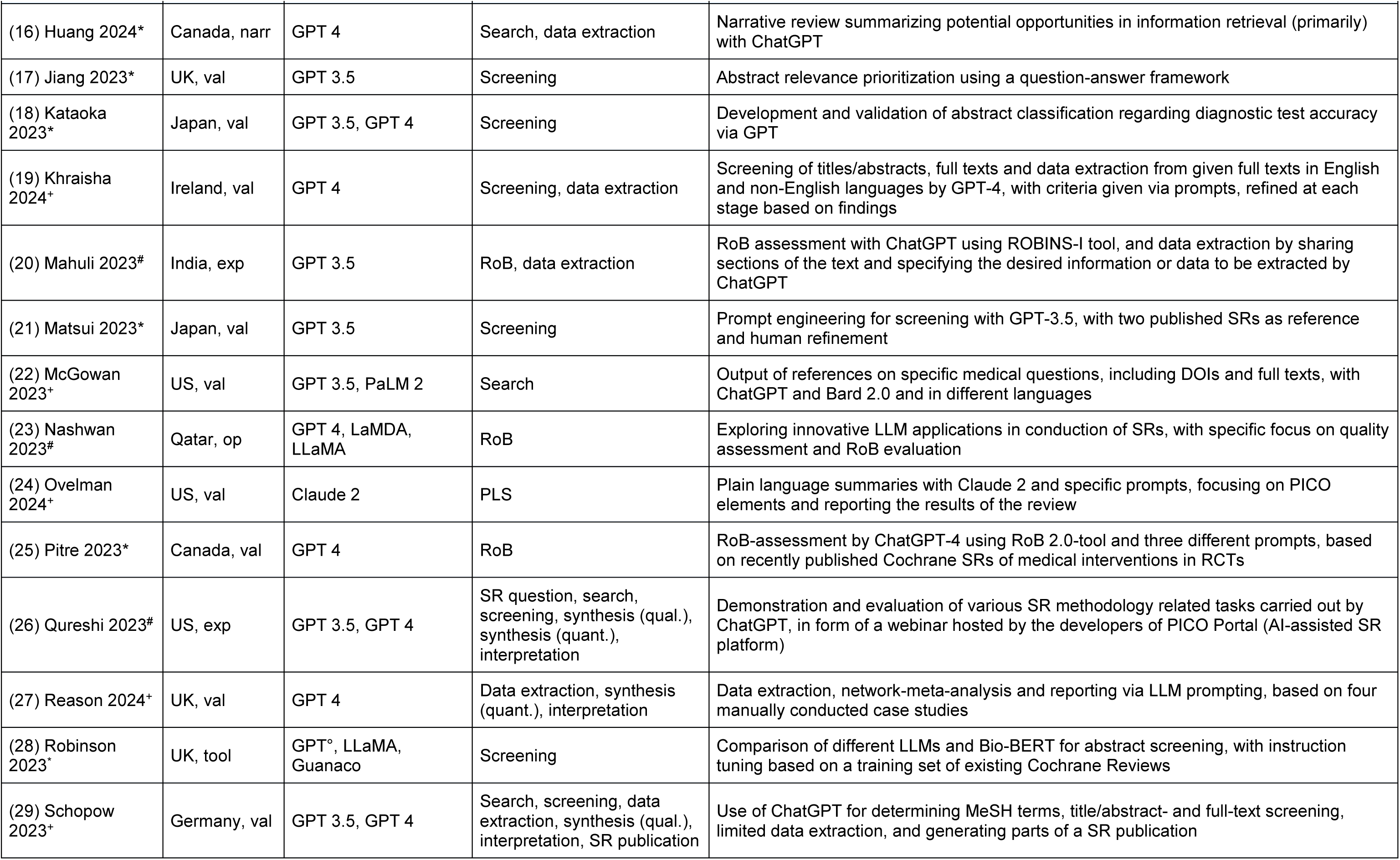
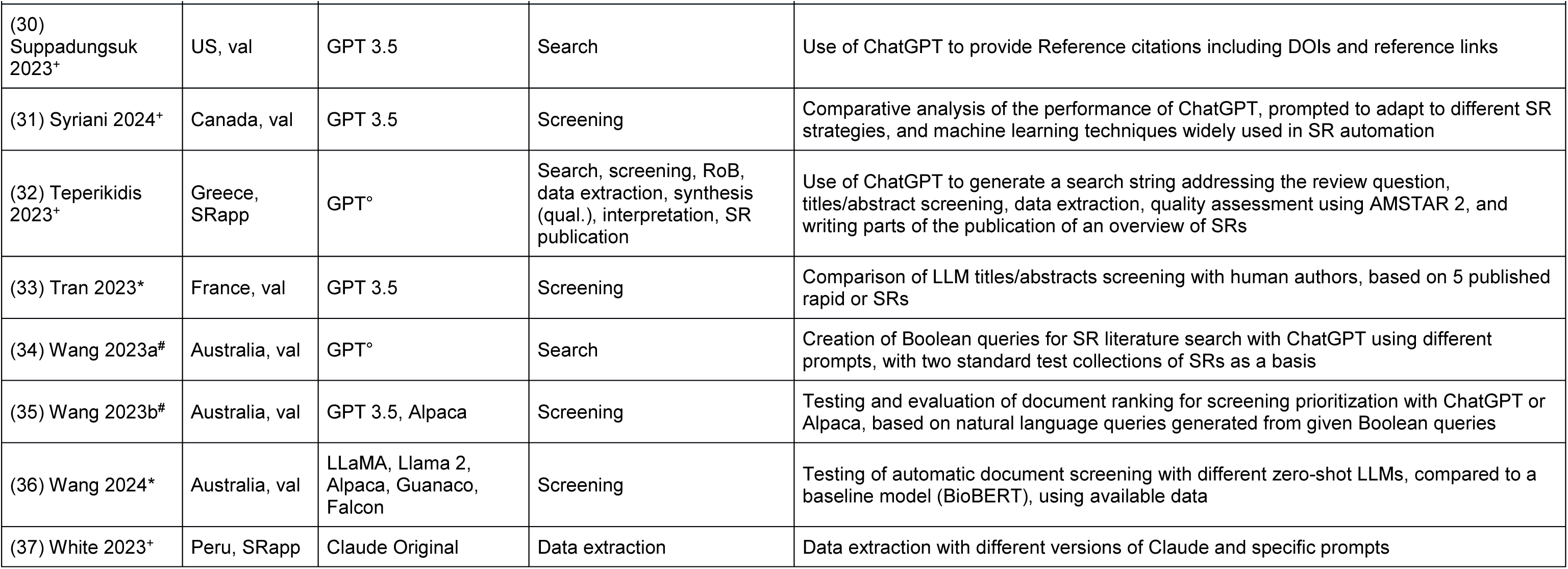
Overview and characteristics of the included studies with LLM applications to support conducting systematic reviews. * preprint, ^+^ journal article, ^#^ communications (comments/letters/editorials/conference proceedings), ^1^ location of the primary institution of the first author, US: United States of America, exp: exploratory testing, val: validation study or accuracy testing, tool: tool development, narr: narrative review, op: opinion, SRapp: LLM application in a systematic review, ° version not specified, RoB: risk of bias assessment, PLS: plain language summary, qual.: qualitative, quant.: quantitative.

In terms of LLMs reported, GPT was the tool used by far most commonly in the studies (n=33, 89% of studies), followed by LLaMA (n=3, 8%), and Claude (n=3, 8%). Details regarding the kind of LLM used for each SR step are depicted in Figure 2 (middle donut) and listed in table 1, including LLM subclasses.

**Figure 2:**
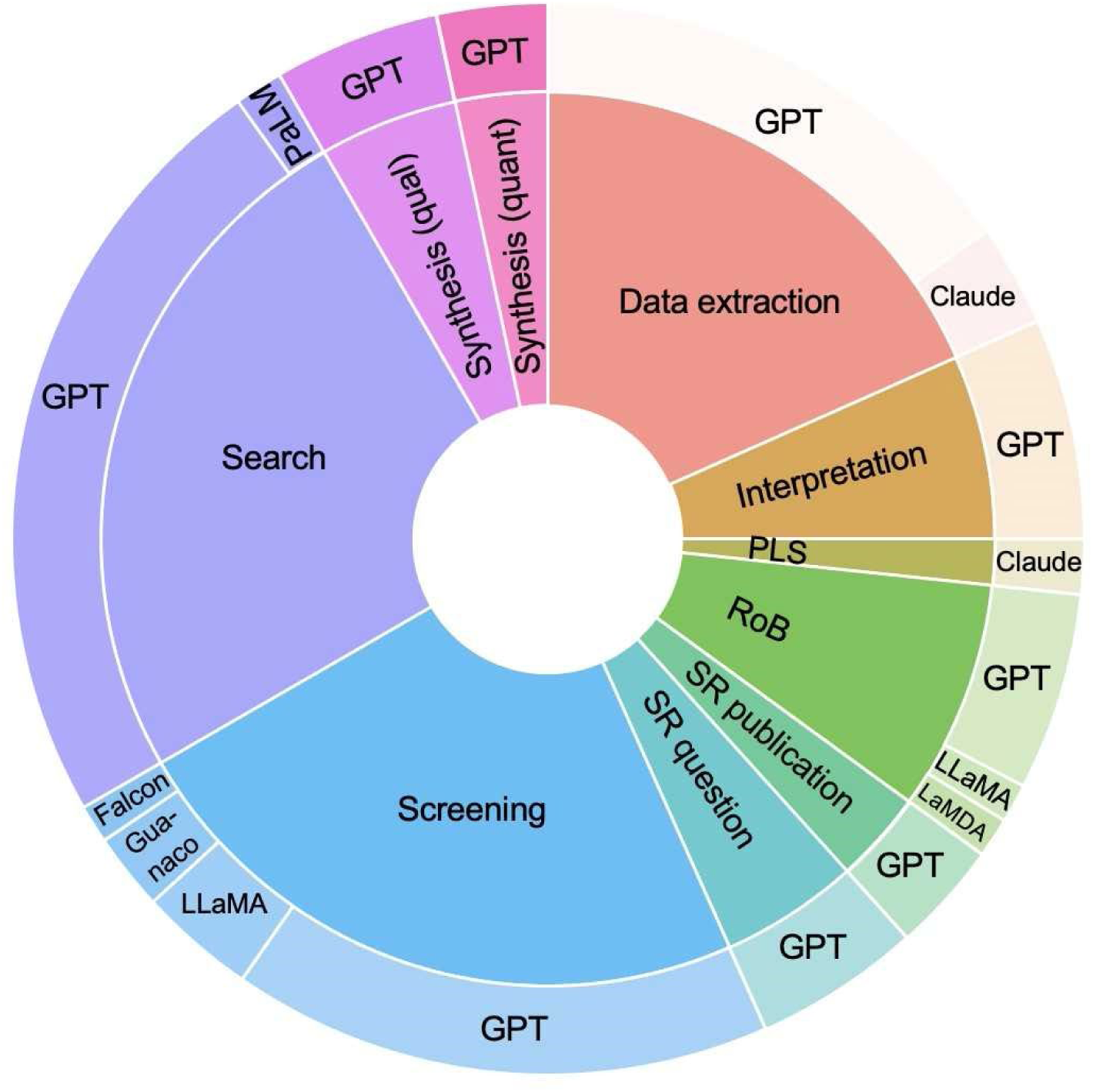
Pie-donut chart depicting proportions of systematic review steps (inner layer pie) and associated LLM applications (outer layer donut). The inner layer represents the frequencies and proportions of all individual SR steps (n=60) extracted across the 37 studies included (multiple counts per study were possible); the outer layer provides a breakdown of the percentage distribution of LLM types used for each SR step: Search (25% of all 60 individual SR steps, n=15. GPT: 100%); Screening (23.3%, n=14. GPT: 70%, LlaMA: 15%); Data extraction (18.3%, n=11. GPT: 83.3%, Claude: 16.7%); RoB (risk of bias assessment; 8.33%, n=5. GPT: 71.43%, LaMDA: 14.3%, LlaMA: 14.3%); Interpretation (6.7%, n=4. GPT: 100%); SR question (5%, n=3. GPT: 100%); Synthesis (qualitative) (5%, n=3. GPT: 100%); Synthesis (quantitative) (3.3%, n=2. GPT: 100%); SR publication (3.3%, n=2. GPT: 100%); PLS (plain language summary; 1.7%, n=1. Claude: 100%).

For half of the studies (n=20, 54%), authors drew a promising conclusion for LLM application in SR conduction, whilst about one quarter of authors concluded neutrally (n=9, 24%), and 22% (n=8) of our 37 reports on LLMs discussed LLM use as non-promising. For example, study prioritization in abstract screening was simplified using a question-answer framework in GPT, and showed higher precision and a substantial increase in efficiency, compared to other zero-shot ranking and BERT-family models [33]. Of the reports of promising findings, screening (n=11) and data extraction (n=6) appeared most often. On the other hand, an approach to evaluate reliability of GPT for performing RoB assessment of randomized trials (RCTs) using the RoB 2.0-tool found only slight to fair agreement between GPT and human reviewers [34]. Supporting the literature search was the SR step that was most frequently reported as non-promising (n=8). Authors’ overall conclusion regarding applicability of LLMs for the respective SR step is summarized in Figure 3.

**Figure 3:**
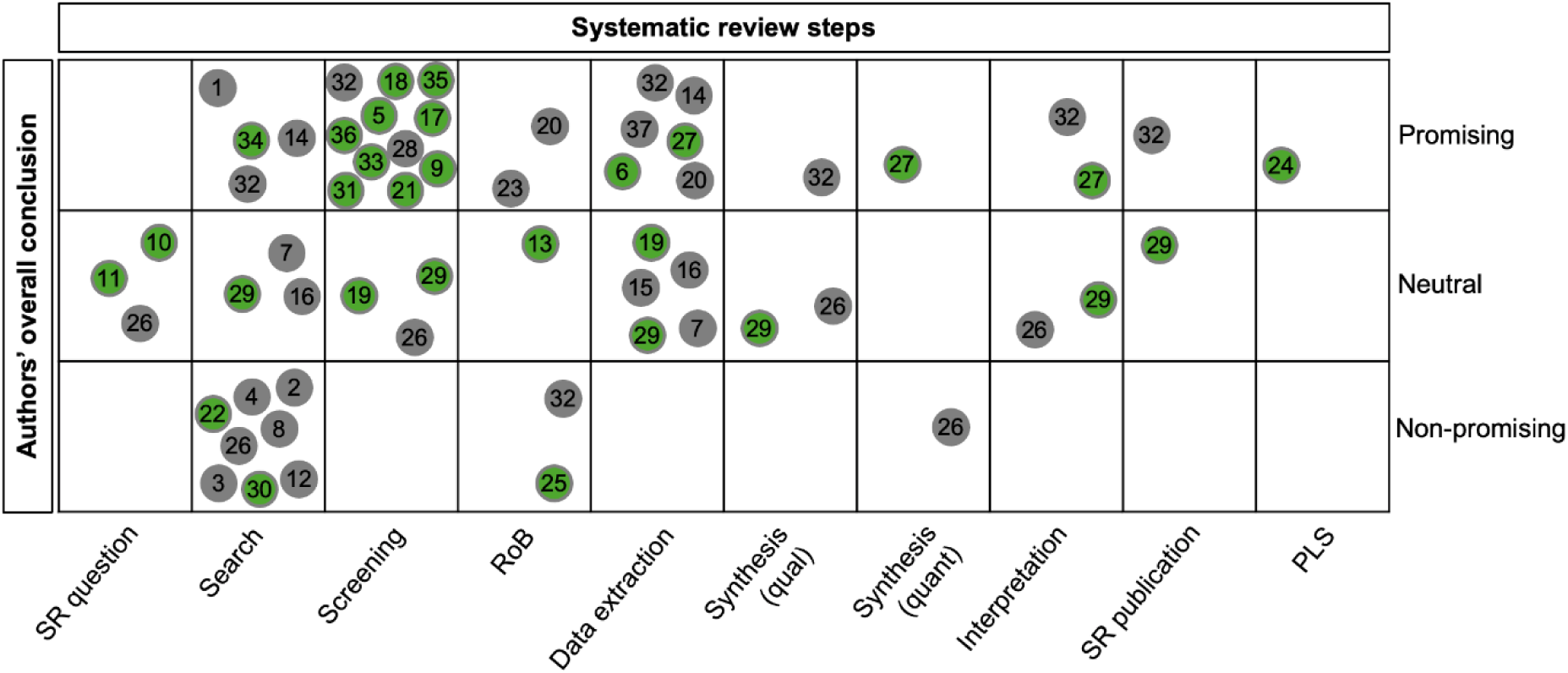
Bubble chart visualizing primary study design (green color: validation studies, grey color: other study designs) and authors’ overall categorized conclusion (y-axis) of each SR step (x-axis). Each bubble represents a study with study-ID as listed in Table 1. Studies evaluating several SR steps are represented multiple times accordingly.

As for study design, the majority of studies were designed as validation studies (n=21, 57%), such as data extraction by Claude compared to human data extraction with already published studies as reference [35], or described an exploratory approach of LLM use for facilitating specific SR steps (n=10, 27%), like formulation of a PubMed search string with GPT [36]. Few authors reported on specific tools they had developed (n=2), such as search engine “Meta-Phill”: Based on PICO components, literature metadata can be saved to a repository on a daily basis, technically supported by ChatGPT API [37]. Two studies were conceptualized as a SR on a medical question but supported by LLM, for example by asking GPT to write parts of the publication [38]. One report gave an opinion on the potentials of LLM application specifically for quality assessment and RoB evaluation in form of an editorial [39]. One article was a narrative review summarizing potential opportunities in information retrieval primarily with ChatGPT [40].

Whilst 70% (n=26) of studies were published in 2023, 30% (n=11) were published in 2024, up to our search date on 26 February 2024. As for type of source, with the reported update in September 2024 on publication status of studies initially published as non-peer-reviewed preprint at the time of our search, 41% (n=15) of the articles were peer-reviewed journal articles, 32% (n=12) were comments, letters, editorials or conference papers, and 27% (n=10) were non-peer-reviewed preprints. Most studies were carried out in the United States (n=11, 30%), followed by the United Kingdom (n=4, 11%) and Canada (n=4, 11%).

## DISCUSSION

In this scoping review, we provide an overview of opportunities and limitations of ML techniques to support SR conduct: We identified 37 articles specifically on LLM applications, where systematic literature search, study selection (screening), and data extraction were most frequent, with OpenAI’s GPT [11] being the most common LLM. Authors evaluated half of the applications as promising and half as neutral to non-promising, highlighting both potential and limitations.

Building on a 2021 mapping review by Jimenez et al. [4], who identified various ML applications in SR performance but no LLMs, we now observed that LLMs have indeed already found their way into many aspects of evidence synthesis: Similar to our LLM findings, Jimenez et al. identified screening, search, and data extraction as the most common ML-supported SR steps. Correspondingly, (semi-) automated title abstract screening and literature search [41] as well as data extraction [42] were the SR steps of greatest significance in further reviews on software and ML support in SRs. However, whilst the majority of publications included in Schmidt et al. [42] describe NLP supported data extraction from abstracts but rarely full texts, we now identified a report on LLM techniques that describes accurate extraction of data presented in text, figures or tables [23]. Contrariwise to training-intensive traditional ML, new LLM approaches require initial prompt engineering but no specific user training [23]. As such, LLMs demonstrated versatility across 10 out of 13 supported SR steps.

Despite the apparent technical simplicity of implementing LLMs in literature search, their evaluation – authors rated half of the approaches as non-promising – shows limitations. In contrast, LLM support in study selection and data extraction appeared more favorable, with by far most (study selection) or at least a slight majority (data extraction) of the described application forms rated as promising, and the rest categorized as neutral.

We revealed notable gaps in LLM use for deduplication, full text retrieval and evaluation of the certainty of evidence using the GRADE approach [43], underscoring constraints. This may be due to their complexity, technical challenges and hence omission of these steps in existing studies, or sufficiency of existing ML tools: Of note, traditional ML tools are already available for the three tasks, such as Deduklick [44] for deduplication, the review tool Covidence [30], which has recently gained some ability for full-text retrieval, and EvidenceGRADEr [45, 46] for evaluating the certainty of evidence.

Regarding the use of LLMs to support SR tasks, there are a number of aspects to keep in mind. LLMs predict the next word based on previous words, resulting in coherent and fluent text output [14]. This creativity aids in tasks like conceiving SR ideas [47] or writing SR reports [38]. Conversely, unlike ML tools like RobotReviewer [48], LLMs do not rely on specialized algorithms and are not originally designed to process structured metadata or ensure transparent reproducibility, which is crucial for high quality scientific literature [49] and evidence synthesis [50]. LLM output greatly varies with temperature settings and the exact wording of the prompt. Lower temperatures result in a more predictable output, higher temperatures enhance diversity in creation [51]. Moreover, minor prompt variations can alter the results [52]. Overall promising GPT-based title abstract screening lacked strict reproducibility even with zero temperature settings [53]. Efforts are underway to balance creativity and coherence in LLM outputs [54]. Capabilities vary depending on input length and session history [55] [56]. Confabulations (“hallucinations”) [57] should be kept in mind.

Further points of criticism include a lack of referencing appropriate and verifiable sources or retrieval of existing literature as well as a variability in non-deterministic LLM responses [21]. Cut-off dates for many LLM training datasets lead to incomplete information [36]. Specifically for validation studies, potential bias due to data contamination must be kept in mind, i.e. if existing studies used as validation reference have already been included into a LLM’s training data set [58]. A way to counteract would be validation studies that use only the newest studies that cannot yet have been included into LLM training datasets. This, however, would significantly reduce the number of studies that can be taken for reference. Because of the authoritative-sounding but potentially inaccurate, incomplete, or biased outcome, some authors suggest application of LLM technology under human supervision and control only; further careful review and editing of the results by authors are needed [59]. The high proportion of validation studies suggests reliability as a basis for future usage, but also the authors’ urge for validation.

Focusing predominantly on GPT calls for caution as opaque decision-making and frequent updates may affect reproducibility [60] and objective and reliable (evidence) research. Other LLMs, such as Claude, offer features particularly suited for SR tasks, including larger context windows and lower hallucination rates [13] [61, 62], rendering a broader selection of LLMs desirable.

Our systematic search reveals a rapid increase in LLM-related publications, including a high proportion of non-peer-reviewed preprints, indicating immense interest in and potential impact for LLMs in SRs. While the sheer volume of papers on the topic may overwhelm peer review procedures, some reports lack sufficient data or methodology. For example, details on prompts were missing in articles [38], or ROBINS-I, conceptualized for RoB assessment of non-randomized studies of intervention, was used to assess RCTs with LLM assistance [63], where RoB 2.0 would have been the correct tool to test. More high-quality validation studies, conducted with strong methodological rigor, are needed for the most promising approaches.

Cierco Jimenez et al. [4] suggest that ML and software developers collaborate to improve already available applications. Similarly, our review can provide a basis for other researchers and app developers in the field.

### Strengths and limitations

This scoping review provides a broad overview of current opportunities and pitfalls in using LLMs in SRs, with no similar work to date. Given the rapidly evolving nature of the field, our review may not be comprehensive, despite thorough searching. New performance assessments for LLM screening [64] [22], data extraction [65], or RoB assessment [24] have emerged since our search. While we aimed for optimal quality in elaborating authors’ overall conclusion as “promising”, “neutral” or “non-promising”, some subjectivity is inherent. As a scoping review, we did not conduct a critical appraisal of data bias. To maintain the highest possible data quality, we followed a strict preregistered protocol and screened a large number of records including those on more traditional ML and BERT approaches. Most of these limitations are characteristic features of a scoping review, giving an orientation on the scope of a heterogeneous topic as a basis for future research.

### Implications for research and practice

Future studies should improve transparency of reporting and ensure a rigorous methodology. Despite the outlined promising aspects, we emphasize the currently still numerous relevant limitations of LLM use, to preserve high quality and unbiased scientific research and evidence synthesis. Nonetheless, we assume that LLMs will play an increasingly important role in SR creation in the future – hopefully backed up with sound research.

## CONCLUSION

This scoping review highlights the rapidly increasing role of LLMs in assisting SR conduction. Albeit promising results in many SR steps, limitations like uncertain reproducibility remain. The surge in publications, including preprints, displays the strong interest and rapid development in the field.

In conclusion, despite in many cases promising, LLMs should currently be used with caution and limited to specific SR tasks under human supervision.

## Supporting information

Supplement_Lieberum2024

## Data Availability

Our preregistered study protocol and extracted LLM and ML data as Excel spreadsheets can be found on OSF (https://osf.io/vdsgb). Supplement material includes our final search strategy and deviations from the preregistered study protocol.

## ABBREVIATIONS AND GLOSSARY

BART: bidirectional and auto regressive transformers
BERT: bidirectional encoder representations from transfomers
DEST-Eppi-Vis: digital evidence synthesis tool evaluations
EBM: evidence-based medicine
GPT: generative pretrained transformer
GRADE: grading of recommendations, assessment, development and evaluation
JBI: Joanna Briggs Institute
KNN: K-nearest neighbors
LaMDA: language model for dialogue applications
LLaMA: large language model Meta AI
LLM: large language model
ML: machine learning
NLP: natural language processing
OSF: open science framework
PaLM: pathway language models
PCC: population, concept, context
PLS: plain language summary
PRISMA-ScR: preferred reporting items for systematic reviews and meta-analyses extension for scoping reviews
RoB: risk of bias
SR: systematic review

## ETHICS STATEMENT

We used secondary data of full scientific articles published in journals or preprint servers. This article does not contain any studies with human or animal participants. Therefore, informed consent and ethical approval were not required.

## ACKNOWLEDGEMENTS

We would like to thank Kathrin Grummich for her advice in developing the draft search strategy, Philipp Kapp for his technical support in the use of Covidence software, Jacqueline Beck for supporting full text retrieval.

## AUTHOR CONTRIBUTIONS

Conceptualization: WS, CH, DB, JJM, AEM

Data curation: JLL, MT, MIM, AEM

Formal analysis: JLL, AEM

Funding acquisition: AEM

Investigation: JLL, MT, MIM, FH, DB, AEM

Methodology: JLL, DB, JJM, AEM

Project administration: JLL, AEM

Resources: DB, JJM

Software: n.a.

Supervision: DB, JJM, AEM

Validation: n.a.

Visualization: JLL, DB

Writing – original draft: JLL

Writing – review and editing: MT, MIM, FH, WS, CH, DB, JJM, AEM

