## Supplement_Lieberum2024 for "Large language models for conducting systematic reviews: on the rise, but not yet ready for use – a scoping review"

### SUPPLEMENTARY MATERIAL

#### I. Detailed search strategies

##### MEDLINE ALL (Ovid)

1. Systematic reviews as topic/
2. (systematic review* or systematic literature review* or scoping review* or rapid review*).tw,kf.
3. (evidence syntheses or evidence synthesis or evidence review* or evidence map* or evidence summar*).tw,kf.
4. or/1-3
5. (artificial intelligence or generative AI or GenAI).ti,kf.
6. (AlexaTM or (Amazon* and Alexa) or Anthropic or Bard or Bardeen or BERT or "Bing chat" or BioGPT or BLOOM or BloombergGPT or Cerebras-GPT or ChatGPT* or "Chat GPT" or chatbot* or Chatsonic or Chinchilla or Claude or DALL-E or EinsteinGPT or Ernie or Falcon or Galactica or "Generative Fill" or "GitHub Copilot" or GLaM or "Google* Assistant" or "Google* Bard" or Gopher or GPT-1 or GPT-2 or GPT-3* or GPT-4* or GPTNeo or GPT-NEoX or GPT-J* or "IBM Watson" or LaMDA or LLaMA or "Megatron-Turing NLG" or "Microsoft* Bing" or Midjourney or Minerva or NeevaAI or Nvidia or OpenAI or "Open AI" or OpenAssistant or OPT or PaLM or PanGu-E or PathAI or "Path AI" or Perplexity or "pre-trained transformer*" or "pretrained transformer*" or (Apple* and Siri) or SlackGPT or StyleGAN or Synthesia or XLNet or YaLM 100B or YouChat).ti,kf.
7. (large language model*).ti,kf.
8. (machine learning or deep learning).ti,kf.
9. (natural language processing).ti,kf.
10. (tool or tools).ti.
11. (automat*).ti,kf.
12. (classifier*).ti,kf.
13. (text mining).ti,kf.
14. or/5-13
15. 4 and 14
16. (202104* or 202105* or 202106* or 202107* or 202108* or 202109* or 20211* or 2022* or 2023* or 2024*).dt.
17. 15 and 16

##### Web of Science Core Collection (SCI, SSCI, ESCI)

#1
TI=("systematic review*" or "systematic literature review*" or "scoping review*" or "rapid review*" or "evidence syntheses" or "evidence synthesis" or "evidence review*" or "evidence map*" or "evidence summar*") OR AB=("systematic review*" or "systematic literature review*" or "scoping review*" or "rapid review*" or "evidence syntheses" or "evidence synthesis" or "evidence review*" or "evidence map*" or "evidence summar*")

#2
TI=("artificial intelligence" or "generative AI" or GenAI or AlexaTM or (Amazon* and Alexa) or Anthropic or Bard or Bardeen or BERT or "Bing chat" or BioGPT or BLOOM or BloombergGPT or "Cerebras-GPT" or ChatGPT* or "Chat GPT" or chatbot* or Chatsonic or Chinchilla or Claude or "DALL-E" or EinsteinGPT or Ernie or Falcon or Galactica or "Generative Fill" or "GitHub Copilot" or GLaM or "Google* Assistant" or "Google* Bard" or Gopher or "GPT-1" or "GPT-2" or "GPT-3*" or "GPT-4*" or GPTNeo or "GPT-NEoX" or "GPT-

J*" or "IBM Watson" or LaMDA or LLaMA or "Megatron-Turing NLG" or "Microsoft* Bing" or Midjourney or Minerva or NeevaAI or Nvidia or OpenAI or "Open AI" or OpenAssistant or OPT or PaLM or "PanGu-E" or PathAI or "Path AI" or Perplexity or "pre-trained transformer*" or "pretrained transformer*" or (Apple* and Siri) or SlackGPT or StyleGAN or Synthesia or XLNet or "YaLM 100B" or YouChat or "large language model*" or "machine learning" or "deep learning" or "natural language processing" or tool or tools or automat* or classifier* or "text mining")

#3
#1 AND #2

#4
Index date: (2021-04-01 to 2024-12-31)

**IEEEXplore**
("systematic review" OR "systematic reviews" OR "systematic literature review" OR "systematic literature reviews" OR "scoping review" OR "scoping reviews" OR "rapid review" OR "rapid reviews" OR "evidence syntheses" OR "evidence synthesis" OR "evidence review" OR "evidence reviews" OR "evidence map" OR "evidence maps" OR "evidence summary" OR "evidence summaries") AND ("artificial intelligence" OR "generative AI" OR GenAI OR AlexaTM OR (Amazon* AND Alexa) OR Anthropic OR Bard OR Bardeen OR BERT OR "Bing chat" OR BioGPT OR BLOOM OR BloombergGPT OR "Cerebras-GPT" OR ChatGPT* OR "GPT" OR chatbot* OR Chatsonic OR Chinchilla OR Claude OR "DALL-E" OR EinsteinGPT OR Ernie OR Falcon OR Galactica OR "Generative Fill" OR "GitHub Copilot" OR GLaM OR "Google Assistant" OR "Google Bard" OR Gopher OR GPTNeo OR "IBM Watson" OR LaMDA OR LLaMA OR "Megatron-Turing NLG" OR "Microsoft Bing" OR Midjourney OR Minerva OR NeevaAI OR Nvidia OR OpenAI OR "Open AI" OR OpenAssistant OR OPT OR PaLM OR "PanGu-E" OR PathAI OR "Path AI" OR Perplexity OR "pre-trained transformer" OR "pre-trained transformers" OR "pretrained transformer" OR "pretrained transformers" OR (Apple* AND Siri) OR SlackGPT OR StyleGAN OR Synthesia OR XLNet OR "YaLM 100B" OR YouChat OR "large language model" OR "large language models" OR "machine learning" OR "deep learning" OR "natural language processing" OR tool OR tools OR automat* OR classifier* OR "text mining")
+ Year Range: 2021 to 2024 + Journals

**Association for Computing Machinery (ACM) Digital Library (nur Proceedings + Journals)**
("systematic review" OR "systematic reviews" OR "systematic literature review" OR "systematic literature reviews" OR "scoping review" OR "scoping reviews" OR "rapid review" OR "rapid reviews" OR "evidence syntheses" OR "evidence synthesis" OR "evidence review" OR "evidence reviews" OR "evidence map" OR "evidence maps" OR "evidence summary" OR "evidence summaries") AND ("artificial intelligence" OR "generative AI" OR GenAI OR AlexaTM OR (Amazon* AND Alexa) OR Anthropic OR Bard OR Bardeen OR BERT OR "Bing chat" OR BioGPT OR BLOOM OR BloombergGPT OR "Cerebras-GPT" OR ChatGPT* OR "GPT" OR chatbot* OR Chatsonic OR Chinchilla OR Claude OR "DALL-E" OR EinsteinGPT OR Ernie OR Falcon OR Galactica OR "Generative Fill" OR "GitHub Copilot" OR GLaM OR "Google Assistant" OR "Google Bard" OR Gopher OR GPTNeo OR "IBM Watson" OR LaMDA OR LLaMA OR "Megatron-Turing NLG" OR "Microsoft Bing" OR Midjourney OR Minerva OR NeevaAI OR Nvidia OR OpenAI OR "Open AI" OR OpenAssistant OR OPT OR PaLM OR "PanGu-E" OR PathAI OR "Path AI" OR Perplexity OR "pre-trained transformer" OR "pre-trained transformers" OR "pretrained transformer" OR "pretrained transformers" OR (Apple* AND Siri) OR SlackGPT OR StyleGAN OR Synthesia

OR XLNet OR "YaLM 100B" OR YouChat OR "large language model" OR "large language models" OR "machine learning" OR "deep learning" OR "natural language processing" OR tool OR tools OR automat* OR classifier* OR "text mining")
+ Year Range: 2021 to 2024 + Publication Type: Journals

**Europe PMC**
TITLE_ABS:("systematic review" OR "systematic reviews" OR "systematic literature review" OR "systematic literature reviews" OR "scoping review" OR "scoping reviews" OR "rapid review" OR "rapid reviews" OR "evidence syntheses" OR "evidence synthesis" OR "evidence review" OR "evidence reviews" OR "evidence map" OR "evidence maps" OR "evidence summary" OR "evidence summaries") AND TITLE:("artificial intelligence" OR "generative AI" OR GenAI OR AlexaTM OR (Amazon* AND Alexa) OR Anthropic OR Bard OR Bardeen OR BERT OR "Bing chat" OR BioGPT OR BLOOM OR BloombergGPT OR "Cerebras-GPT" OR ChatGPT* OR "GPT" OR chatbot* OR Chatsonic OR Chinchilla OR Claude OR "DALL-E" OR EinsteinGPT OR Ernie OR Falcon OR Galactica OR "Generative Fill" OR "GitHub Copilot" OR GLaM OR "Google Assistant" OR "Google Bard" OR Gopher OR GPTNeo OR "IBM Watson" OR LaMDA OR LLaMA OR "Megatron-Turing NLG" OR "Microsoft Bing" OR Midjourney OR Minerva OR NeevaAI OR Nvidia OR OpenAI OR "Open AI" OR OpenAssistant OR OPT OR PaLM OR "PanGu-E" OR PathAI OR "Path AI" OR Perplexity OR "pre-trained transformer" OR "pre-trained transformers" OR "pretrained transformer" OR "pretrained transformers" OR (Apple* AND Siri) OR SlackGPT OR StyleGAN OR Synthesia OR XLNet OR "YaLM 100B" OR YouChat OR "large language model" OR "large language models" OR "machine learning" OR "deep learning" OR "natural language processing" OR tool OR tools OR automat* OR classifier* OR "text mining")
+ Year Range: 2021 to 2024 + Preprints

**Google Scholar**
("systematic reviews" | "systematic literature reviews" | "scoping reviews" | "rapid reviews" | "evidence syntheses" | "evidence synthesis" | "evidence reviews") ("artificial intelligence" | "generative AI" | genAI | ChatGPT | GPT | "large language model" | "large language models" | "machine learning" | "deep learning" | "natural language processing" | tool | tools | automation | automated | automate | classifier | classifiers | "text mining")
Filter: 2021 to 2024 / ranked by relevance / downloaded first 200 hits

### II. Deviations from the protocol

Contrary to our protocol, we

- did not perform an additional search in Cochrane Evidence Synthesis and Methods (Wiley) because we found that the journal was already indexed in MEDLINE and has therefore already been included in our systematic search,
- were not able to search the systematic review toolbox (https://systematicreviewtool.com) because the webpage was not available during the entire time period from the search to the preparation of the manuscript (February to October 2024),
- additionally searched the Digital Evidence Synthesis Tool (DEST) Evaluations [1] of which we were not yet aware at the time of writing the protocol.

[1] Bond M, Finnerty A, O'Mara-Eves A, O'Driscoll P, Thomas J, Minx J, Callaghan M, Scheelbeek P. Digital Evidence Synthesis Tool Evaluations. EPPI Visualiser database. 2024. https://eppi.ioe.ac.uk/eppi-vis/Review/Index/435
